# Neural correlates of β-lactam exposure in intensive care unit patients: an observational, prospective cohort study

**DOI:** 10.1101/2024.09.25.24314366

**Authors:** Arnaud Zalta, Agnès Trébuchon, Géraldine Daquin, Lionel Velly, Marc Leone, Olivier Blin, Stanislas Lagarde, Romain Guilhaumou

## Abstract

**Background:** Β-Lactam-induced neurotoxicity in critical care patients can compromise clinical outcomes. Despite the growing use of therapeutic drug monitoring (TDM) for β-lactams, clear toxicity thresholds remain undefined, leaving clinicians uncertain about dosing adjustments when adverse effects occur. Identifying a relevant and easily detectable neurophysiological biomarker for β-lactam exposure would improve monitoring and prevent serious complications.

**Methods:** In a prospective multicenter, non-interventional study, we analysed electroencephalographic (EEG) signals of 56 patients hospitalized in intensive care units (ICUs) receiving continuous infusions of five β-lactams (meropenem, piperacillin/tazobactam, cefepime, cefotaxime, or ceftazidime). We applied a time frequency decomposition on these EEG data to investigate quantitatively the power of neural dynamics across frequencies ranging from 1 to 45 Hz. We used a multivariate pattern decoding method to correlate the β-lactam exposure and Sepsis-related Organ Failure Assessment (SOFA) scores with the neural activity.

**Results:** β-lactam exposure correlated with increased β-low γ neural dynamics (20–40 Hz) (p < 0.001, FDR corrected), independent of other clinical factors or medications. Β neural activity was most pronounced in central electrodes (C3 : r = 0.20, p < 0.01; C4 : r = 0.26, p < 0.01) and the right frontal electrode (Fp2 : r = 0.12, p = 0.02). Lower θ-α activity (3.5-5 Hz and 12-18 Hz) was associated with higher SOFA scores (p < 0.001, FDR corrected). No significant correlations were observed between other drugs (opioids, antiseizure medications, psychotropics) and β or θ-α dynamics.

**Conclusions:** These results suggest that β neural dynamics represent a potential biomarker for β-lactam exposure in ICU patients. They highlight the potential of quantitative EEG and advanced multivariate decoding methods to identify subtle neurophysiological features that are otherwise difficult to detect.

**Trial registration:** ClinicalTrials.gov ID NCT03339869. Registered 14 September 2017.

## Background

B-lactams are the most widely prescribed antibiotics due to their broad-spectrum activity and relatively good tolerance (1–4). Despite their high benefit-to-risk ratio, they induce adverse events, including gastrointestinal disorders, hypersensitivity reactions, and neurological disorders such as encephalopathy and seizures (5–9).

These adverse events are particularly frequent in vulnerable populations, such as intensive care unit (ICU) patients (10). Neurological injuries are common in ICU patients and can significantly impact clinical outcomes, making them susceptible to the neurotoxic effects of β-lactam antibiotics (11–13). Additionally, the cumulative organ failures in ICU patients lead to high pharmacokinetic variability, further increasing the risk of β-lactam overexposure and induced neurotoxicity (10).

To mitigate these risks, therapeutic drug monitoring (TDM) of β-lactams has become increasingly recommended over the past decade, guiding antimicrobial dosing to optimize therapeutic effects (14,15). However, despite the growing use of TDM, data on the toxic concentrations of β-lactams are still limited, and clear toxicity thresholds have yet to be established for these antibiotics (14,16). As a result, clinicians often face the dilemma of reducing or discontinuing treatment when adverse effects arise (14).

In addition, β-lactams can affect organ function, including brain activity, but these effects are difficult to measure and correlate accurately. Continuous electroencephalographic (EEG) monitoring has identified pathological patterns in patients suffering from β-lactam-induced neurotoxicity, particularly with the use of cefepime. These patterns include generalized rhythmic sharp waves, periodic discharges, and other abnormal waveforms (17). However, these EEG signatures are nonspecific to β-lactam intoxication and require subjective, time-consuming interpretation by physicians (18). In addition, myoclonus and seizures have been noted in patients with antibiotic-associated encephalopathies, which is characteristic of penicillins and cephalosporins (19,20).

Given the limitations of traditional EEG analysis, there is a growing interest in leveraging quantitative EEG (qEEG) analysis to automate and enhance the detection of neurotoxicity. While promising, qEEG has not yet been widely adopted in clinical practice (21). Some studies have shown that β-lactam antibiotics can modulate EEG signals, particularly in the frequency domain, but these studies have largely been conducted in animal models and typically at a single dose of antibiotics (22–24).

In this context, we investigated potential new qEEG markers in 56 ICU patients receiving continuous intravenous β-lactam antibiotics. Our study used multivariate pattern analysis, a powerful method not commonly employed in clinical practice. By applying this technique to the amplitude of frequencies in the EEG signal, we aimed to identify biomarkers that would result in an automated, objective detection of β-lactam exposure. Multivariate pattern analysis enhances the impact of our research by providing a more precise approach to detecting subtle changes in brain activity, which would improve patient safety and optimize therapeutic outcomes without compromising antimicrobial efficacy.

## Methods

### Patients and design

The current analysis was part of a prospective, multicenter trial conducted at two Intensive Care Units (see the Declarations section). Inclusion criteria required patients to be aged 18 or above, expected to stay in the critical care unit for at least seven days, and receiving one of the five specified β-lactam antibiotics (meropenem, piperacillin/tazobactam, cefepime, cefotaxime, or ceftazidime). Exclusion criteria included pregnancy, known hypersensitivity to β-lactam antibiotics, lack of social security affiliation, or inability to obtain consent.

### Plasma β-lactam concentration determination

Samples were collected at the patient’s bedside from a puncture site remote from the antibiotic injection site. Results were typically returned within 24 hours of the assay. Dosage adjustments were made as deemed necessary by the clinician and pharmacologist. The target total steady-state plasma concentrations following continuous administration for cefepime, cefotaxime, ceftazidime, piperacillin/tazobactam, and meropenem were 5-35 µg/mL, 25-60 µg/mL, 40-80 µg/mL, 80-160 µg/mL, and 8-16 µg/mL, respectively. To establish a class relationship between β-lactam exposure and quantitative changes in EEG, plasma concentrations of each antibiotic were normalized by the minimum inhibitory concentrations (MIC). The respective MIC thresholds, selected by considering β-lactam treatment either during the empirical phase or in cases with no microbiological documentation (14), were 1 µg/mL for cefepime, 4 µg/mL for cefotaxime, 8 µg/mL for ceftazidime, 16 µg/mL for piperacillin/tazobactam, and 2 µg/mL for meropenem.

Heparinized tubes containing the samples were transported to the analysis laboratory within 5 hours of collection. The samples were then centrifuged for 10 minutes at 3,000g and 4°C. For assays involving meropenem, a solution of 4-morpholinoethanesulfonic acid was added. Protein precipitation was performed using acetonitrile, followed by liquid-liquid extraction with dichloromethane. An in-house standard, 5-methoxyindole-3-acetic acid, was used for the β-lactam antibiotics. Antibiotic concentrations were measured using high-performance liquid chromatography coupled with ultraviolet detection (Dionex Ultimate 3000 HPLC) (25). The limit of quantification for all antibiotics was 0.5 μg/mL, with a validated upper limit of linearity at 50 μg/mL for meropenem and 100 μg/mL for the other β-lactams. Samples exceeding these limits were diluted and re-analysed. This protocol was adapted from the method described by (26) and complies with recommendations from the European Medicines Agency (27).

### SOFA score determination

The SOFA (Sequential Organ Failure Assessment) score is a scoring system designed to track the progression of organ failure in critically ill patients. It assesses the function of six organ systems :

1. Respiratory: Assessed by the PaO2/FiO2 ratio or the need for mechanical ventilation.
2. Coagulation: Evaluated by measuring platelet count.
3. Liver: Assessed by serum bilirubin levels.
4. Cardiovascular: Assessed using vasopressor agents and mean arterial pressure.
5. Renal: Assessed by serum creatinine levels and urine output.
6. Central Nervous System: Assessed by the Glasgow Coma Scale.

Each organ system is scored from 0 (normal function) to 4 (severe dysfunction), with the total score providing an overall assessment of a patient’s condition. Higher scores indicate greater organ dysfunction and are associated with worse outcomes (28).

### EEG data acquisition

EEG data were recorded using an 11-channel Deltamed^©^ clinical system, sampled at 256 Hz. The electrodes were placed according to the 10/20 system (29). After recording, the raw EEG data were high-pass filtered at 1 Hz using a 4th-order Butterworth filter to remove slow drifts (e.g., sweat-induced) and then referenced to the average of all electrodes. Power line artifacts were removed using notch filters at 50 Hz. Stereotyped artifacts, such as eye blinks and cardio-related signals, were eliminated using Independent Component Analysis. To ensure consistency in signal-to-noise ratio both within and across patients, amplitude normalization was applied using z-scoring across the time dimension for each EEG recording. These preprocessing steps were executed with the FieldTrip toolbox for MATLAB (30) and supplemented with custom scripts (Matlab 2018b, The MathWorks).

### Spectral decomposition of the EEG data

The EEG data were subsequently segmented into one-minute epochs following the time-frequency decomposition of the entire EEG recording. This decomposition was performed using 50 logarithmically spaced frequencies ranging from 1 to 45 Hz. The Morlet wavelet transform was applied to the data using the FieldTrip toolbox (30).

### Multivariate pattern analysis on channel-level EEG data

Multivariate pattern decoding analyses were conducted across patients by capitalizing on the spatial patterns of the EEG power, i.e., spectrally decomposed and time-averaged for each neural frequency and each regressor (β-lactam exposure at Day 4, SOFA score at Day 4). We used a cross-validated multivariate linear decoding model to estimate the spatial EEG patterns *ŵ* of a specific data associated with each regressor characteristic X. For each cross-validation fold (n = 10, interleaved, see (31), we defined the spatial EEG patterns *ŵ* on the training set by regressing—in a ridge sense (ridge α parameter set at 2) —each EEG feature *Z*_*train*_(11 channels in total) against the regressor characteristic *X*_*train*_ across regressor exemplars, by solving 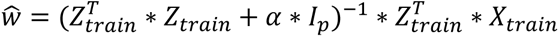, where *I*_*p*_ is the *p*p* identity matrix with *p* corresponding to the number of EEG channels (11 in total). We then projected the EEG data on the test set *Z*_*test*_, on the dimension defined by the coding weights *ŵ* to obtain neural predictions of the regressor characteristic *X*_*test*_ for each epoch of the test set. Note that the training and testing set never include epochs (20 per patient) from a same patient to avoid possible overfitting effect. After applying this procedure for each cross-validation fold, we computed the linear Pearson’s correlation coefficient between neural predictions *X̂* and ground-truth values X of the regressor characteristic. The coding precision metric reported in the main text corresponds to the Fisher transform of the correlation coefficient, which is approximately normally distributed.

Multivariate pattern decoding analysis described above was conducted on orthogonalized regressors using a General Linear Model procedure at the group level.

### Statistical procedures

All analyses were conducted using standard nonparametric tests at the group level, such as the Wilcoxon rank-sum test. Channel-level correlations were assessed using Pearson correlations. To control the type 1 error rate from multiple comparisons, we applied the False Discovery Rate correction across the dimensions of interest, specifically electrodes and frequencies, following the procedure introduced by (32).

## Results

### Population characteristics

From 01 December 2018 to 30 December 2020, 120 ICU patients admitted were prospectively included in the study. The EEG recordings were conducted on the fourth day of inclusion for 72 of these patients. Ultimately, 56 patients (37 males, 66%) with EEG recordings of suitable quality were included in our analysis (Figure 1).

**Figure 1.**
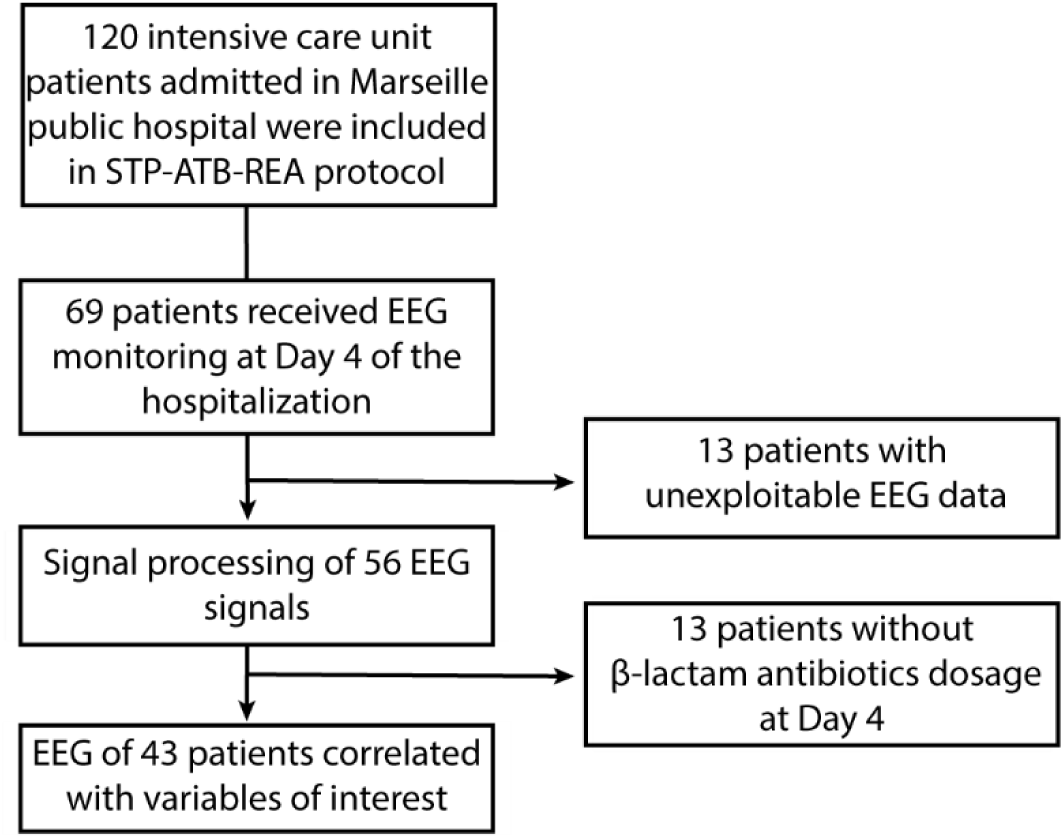
Study flow chart

The average age of the participants was 61 years, with an average weight of 75 kg. On the fourth day of inclusion in the protocol, when β-lactam antibiotics dosage and EEG recordings were performed, the patients had an average creatinine clearance of 122 mL/min and an albumin level of 27 g/L (see table 1 for more details).

**Table 1.**
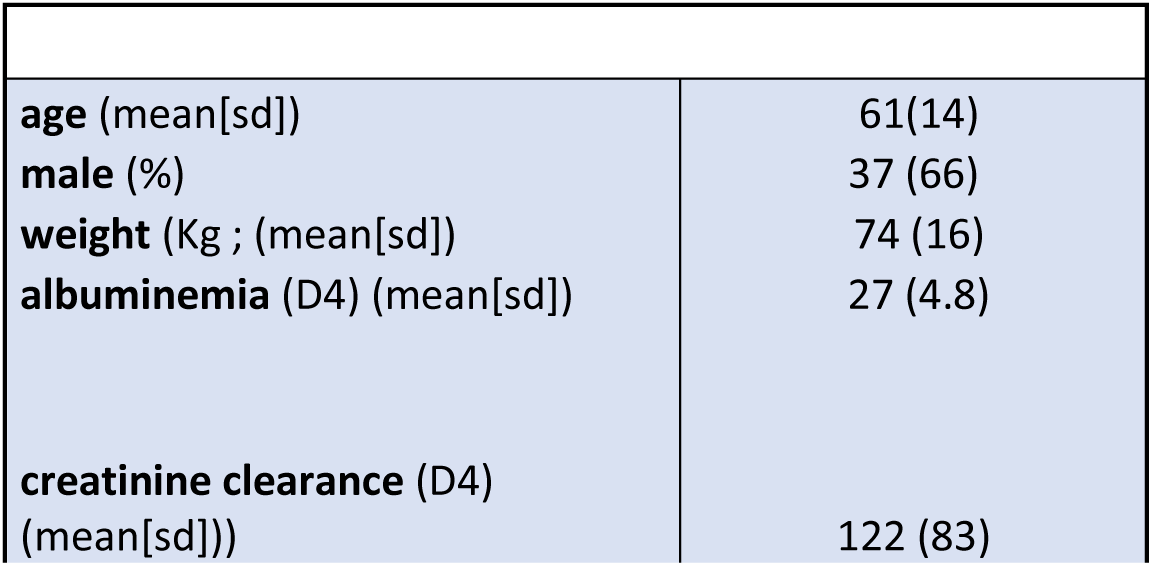
Demographic and clinical characteristics of the population. Standard deviation (sd); kilogram (Kg); value at day four after beginning of hospitalisation (D4).

| **age** (mean[sd]) | 61(14) |
| --- | --- |
| **male** (%) | 37 (66) |
| **weight** (Kg ; (mean[sd]) | 74 (16) |
| **albuminemia** (D4) (mean[sd]) | 27 (4.8) |
| **creatinine clearance** (D4) (mean[sd])) | 122 (83) |

Among the cohort, 31 (55%) patients were admitted to ICU for acute neurological injury, with no reported cases of neurodegenerative pathology: 6 patients with stroke, 24 with intracranial bleeding, 1 with seizures and 14 with traumatic brain injury. 3 patients were admitted for meningitis infection. In addition, 5 cases of a history of chronic alcoholism were reported. The Sepsis-related Organ Failure Assessment (SOFA) score was assessed at 4.7 (SD = 3.1).

### Pharmacological features

At inclusion, the patients received piperacillin/tazobactam (n = 26), cefotaxime (n = 17), meropenem (n = 6), cefepime (n = 4), and ceftazidime (n = 3). On the fourth day of the protocol, 2 patients who had initially started treatment with meropenem were switched to cefotaxime. Plasma β-lactam concentrations were measured for 43 patients on day 4. Of these, 15 (35%) patients had plasma concentrations within the target range, while 25 (58%) patients were underdosed and 3 (7%) patients were overdosed.

To assess β-lactam exposure, we normalized the β-lactam concentration relative to the Minimum Inhibitory Concentration (MIC) of the respective antibiotic for each patient (see Methods). The mean β-lactam exposure was 6.23 (SD = 5.26; arbitrary units).

### Concomitant treatment

We compiled a list of medications that could potentially influence brain activity and affect the EEG recordings on day 4 of the protocol. Among the patients, 24 (43%) received opioids, 7 (13%) antiseizure medications, 3 (5%) ketamine, and 6 (11%) psychotropic drugs. Additionally, 18 patients (32%) were treated with benzodiazepine, and 2 patients (3.6%) received anticancer drugs (Table 2).

**Table 2.** Treatments or drug substances (alcohol, psychotropics) administered before the hospitalization or at ICU admission.

| Concomitant medications | N (%) |
| --- | --- |
| opioids (%) | 24 (43) |
| antiseizure medications (%) | 7 (13) |
| ketamine (%) | 3 (5.4) |
| cancer (%) | 2 (3.6) |
| psychotropes(%) | 6 (11) |
| benzodiazepine (%) | 18 (32) |
| anti-cancer drugs (%) | 2 (3.6) |

### Decoding analyses

To investigate the neural correlates of β-lactam exposure across different EEG frequencies, we conducted multivariate pattern (decoding) analyses on the EEG data at the channel level. Our analysis revealed that β-lactam exposure was encoded in α neural activity between 12.5 and 14.5 Hz and, more notably, in the broad β-low γ band between 20 and 40 Hz (Figure 2a, p < 0.001 FDR corrected).

**Figure 2.**
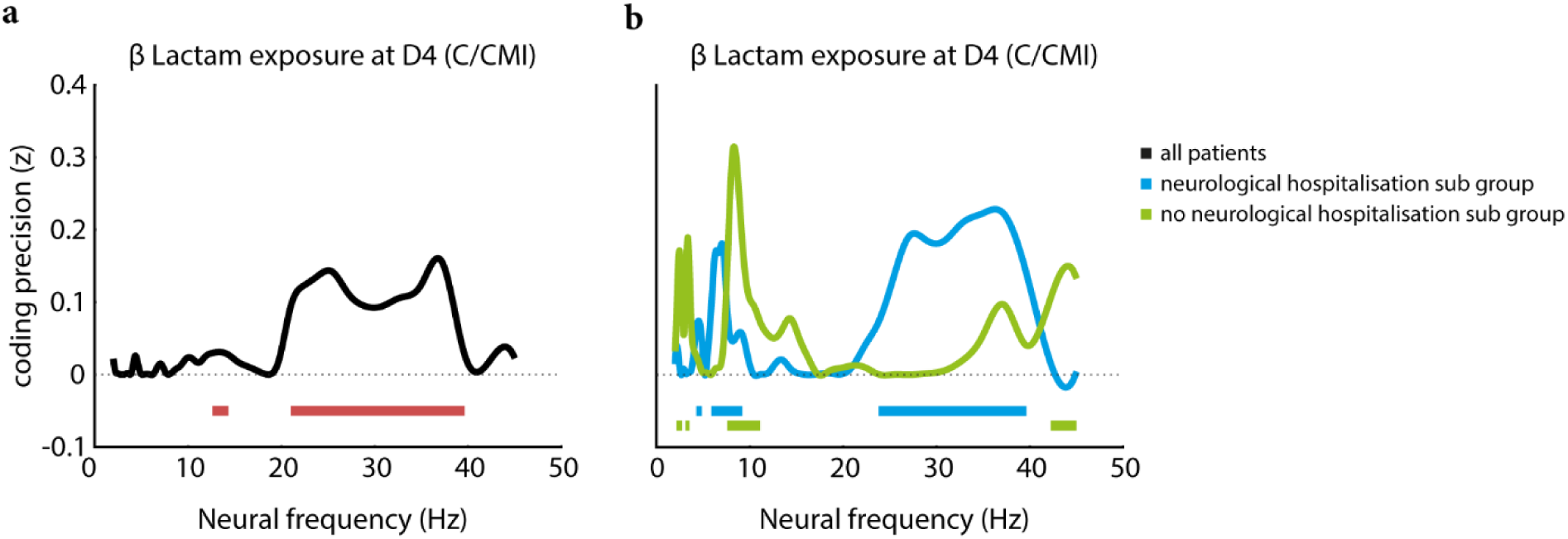
**a**. Spectrum of neural coding of the β-lactam exposure at the fourth day of the hospitalization of the entire population and **b.** for two subgroups: from patients admitted for acute neurological injury (blue line) or not (green line). Red, blue and green horizontal lines indicate frequencies with significant coding values for the entire population, group of patients admitted for acute neurological injury and not, respectively (q < 0.001, FDR-corrected).

Next, we examined the correlation between β-lactam exposure and β (20-40 Hz) neural dynamics at the electrode level. We found a positive correlation between β neural activity and β-lactam exposure in central electrodes (C3: r = 0.20, p < 0.01; C4: r = 0.26, p < 0.01; Figure 3). In addition, a right frontal electrode (Fp2) showed a positive correlation with β-lactam exposure (r = 0.12, p = 0.02; Figure 3). No other electrodes exhibited statistically significant correlations. We observed similar results with a right frontal electrode correlating high θ neural activity with β-lactam exposure (Fp2: r = 0.12, p < 0.01; Supplementary Figure 2).

**Figure 3.**
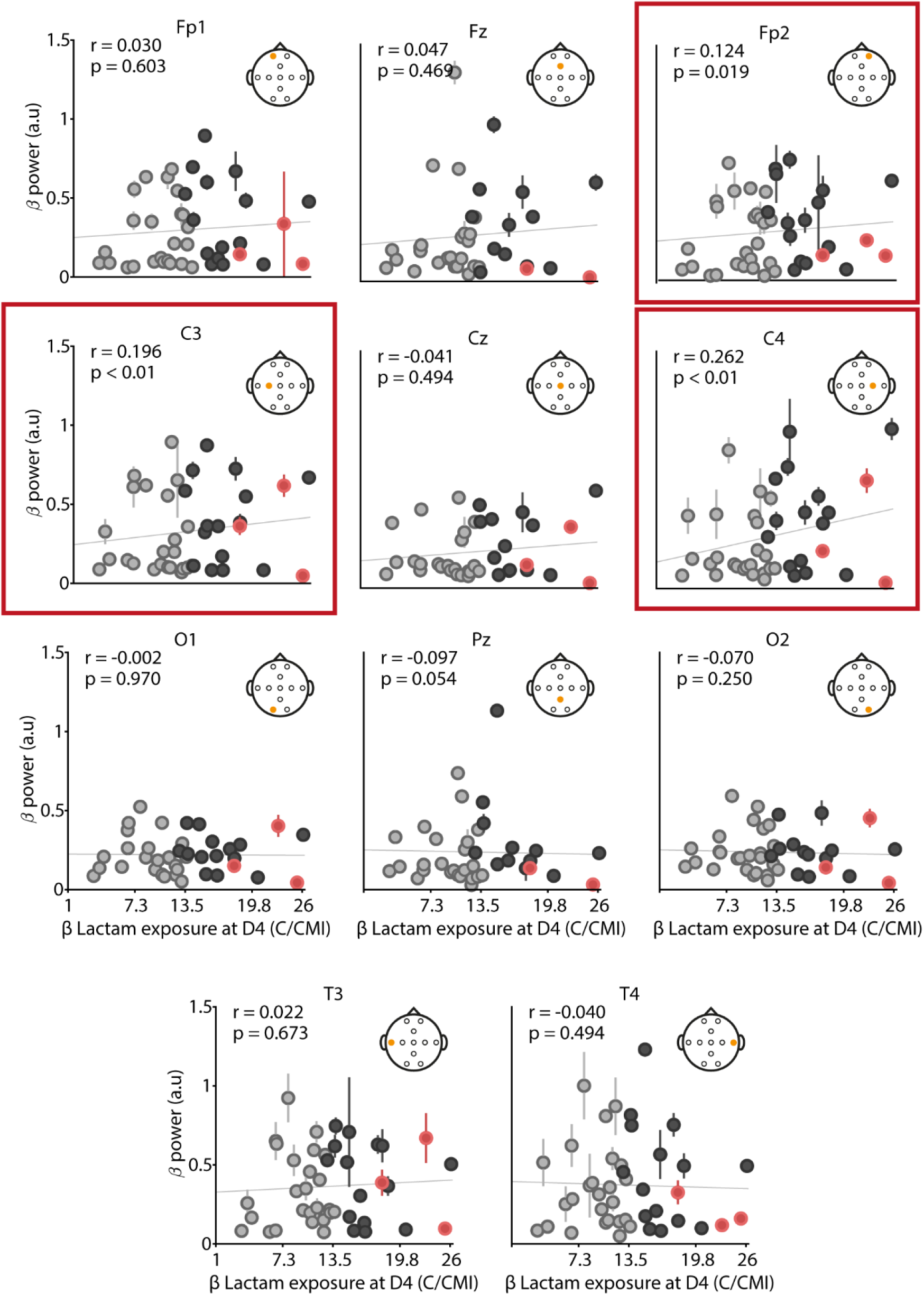
Correlation between the power of β neural dynamics for each EEG electrodes and the β-lactam exposure (concentration (C) over MIC; see methods). Each point relates to one patient. Grey, black and red points correspond to the patients labelled as sub-, well and overdosed respectively. Electrodes outlined in red show a positive correlation statistically significant (p<0.01).

Next, to assess the effect of neurological injury on patients’ EEGs, we performed multivariate pattern analysis, dividing the dataset into two subgroups. The first subgroup comprised patients without acute neurological injury, while the second included patients with acute neurological injury. In the subgroup without acute neurological injury, β-lactam exposure was encoded in the δ (2-3.5 Hz), θ (8-11 Hz), and low γ (42-45 Hz) bands, but not in the previously identified β band (Figure 2b, p < 0.001 FDR corrected). In contrast, in the subgroup with acute neurological injury, β-lactam exposure was encoded in the θ-low α (4-9 Hz) and a broad β-low γ (23-45 Hz) bands (Figure 2b, p < 0.001 FDR corrected). Notably, a band between 7 and 10 Hz was associated with β-lactam exposure in both subgroups (Figure 2b, p < 0.001 FDR corrected). None of the other clinical or pharmacological features significantly affected β or high θ neural dynamics (df = 10.8; p > 0.05; Supplementary Figure 3), except for chronic alcohol consumption, which significantly decreased high θ neural dynamics (presence vs. absence of alcohol consumption, t = −3.35; p < 0.01; Supplementary Figure 4).

Finally, we examined the association of EEG patterns and the SOFA score. The SOFA score correlated with θ (3.5-5 Hz) and α (12-18 Hz) neural dynamics, but not with higher frequencies (Figure 4, p < 0.001 FDR corrected). At the electrode level, we found a significant negative correlation between the SOFA score and α neural dynamics for all electrodes (r < - 0.14, p < 0.01; Figure 5). Like other frequency bands, no other clinical or pharmacological features significantly affected α neural dynamics, except for chronic alcohol consumption, which significantly decreased α neural dynamics (presence vs. absence of alcohol consumption, t = −2.26; p < 0.01). No significant effects were observed for other features (df = 8.96; p > 0.05; Supplementary Figure 5).

**Figure 4.**
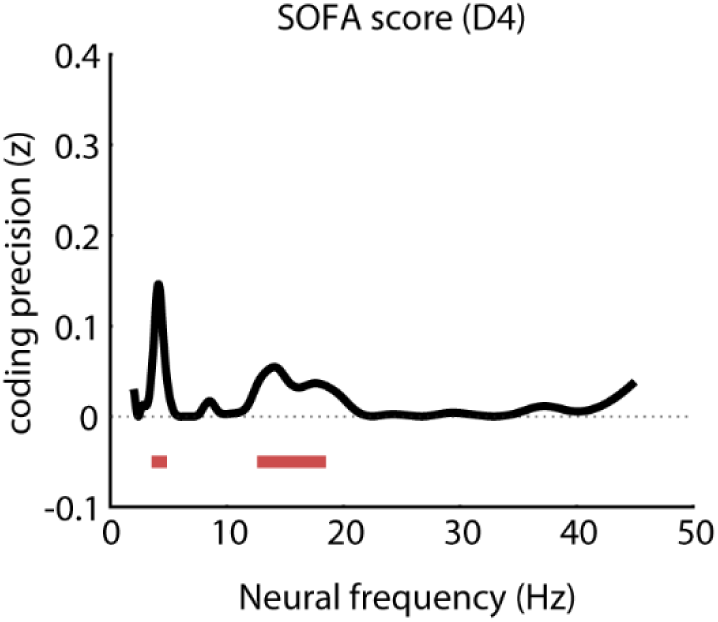
Spectrum of neural coding of the SOFA score computed at the fourth day of the hospitalization of the entire population; from whole electrodes EEG signals. Red horizontal lines indicate frequencies with significant coding values (q < 0.001, FDR-corrected).

**Figure 5.**
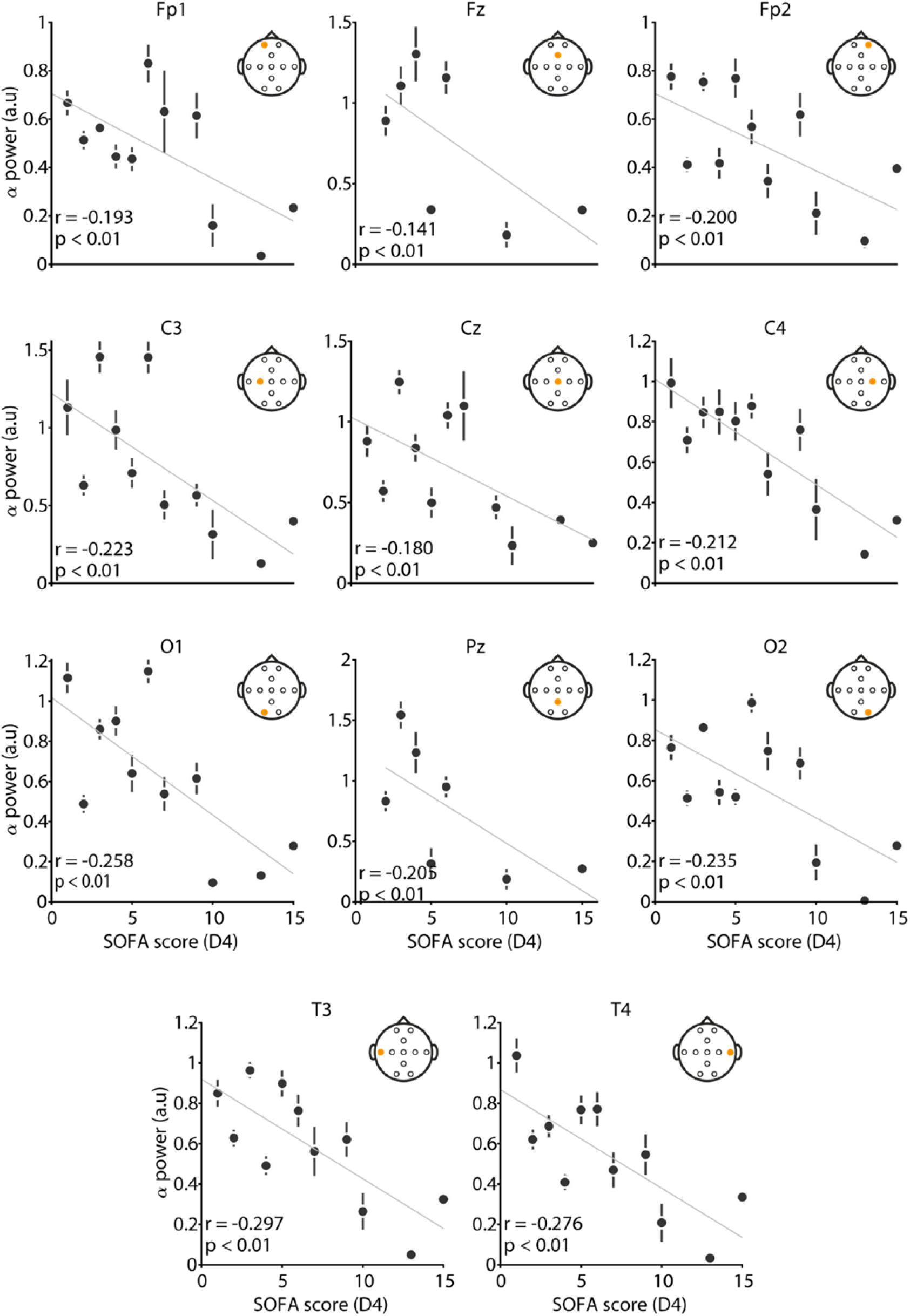
Correlation between the power of the α neural dynamics for each EEG electrode and the SOFA score at day 4 (D4). Each point relates to one level of SOFA score. All Electrodes show statistically significant negative correlation (p<0.01).

## Discussion

The aim of our study was to identify an EEG marker of β-lactam exposure in ICU patients. Using quantitative EEG, we found that the amplitude of neural dynamics in the β/low-γ frequency range (20-40 Hz) correlated with β-lactam exposure, while lower θ-α dynamics code for the SOFA score. This analysis was based on a frequency-specific examination of EEG data, utilizing machine learning to predict β-lactam exposure from the amplitude of β EEG neural dynamics. Univariate correlations across EEG channels did not provide strong evidence in favour of a direct relationship between β-lactam exposure and β EEG dynamics. Our multivariate machine learning approach, which accounts for the spatial dimensions of all EEG sensors, offers a more powerful approach.

Our findings of increased high θ and β neural dynamics associated with β-lactam exposure align with previous studies conducted on anesthetized rats (22,33). These studies suggest that β neural dynamics might indicate neuronal hyperexcitability (34,35). The mechanism behind β-lactam-induced neurotoxicity is thought to involve either non-competitive (e.g., penicillins) or competitive (e.g., cephalosporins) inhibition of type A Gamma Aminobutyric Acid (GABA-A) receptors, thereby impairing inhibitory neurotransmission (36–40). Animal studies have shown that penicillins decrease inhibitory postsynaptic potentials (IPSPs) and increase excitatory neuron burst properties, which may lead to myoclonus and seizures (41–45).

Studies showed that GABA signaling modulation affected β neural dynamics through interactions between cortical and subcortical circuits (46,47). Reports of amoxicillin-induced neurotoxicity with seizures and spike discharges displaying generalized β neural dynamics on EEG further support this hypothesis (23,48). Myoclonus and convulsive seizures associated with type I antibiotic-induced encephalopathies, characteristic of penicillins and cephalosporins, have also been documented (19). Common EEG abnormalities in these cases include nonspecific signs of encephalopathy, such as slowing and generalized periodic discharges with triphasic waves. In cephalosporin-associated encephalopathy, 55% of cases exhibited epileptiform discharges or seizures, while 40% of penicillin-associated cases showed similar findings. Continuous EEG analysis revealed pathological patterns, including generalized rhythmic sharp and slow waves, periodic discharges, acute biphasic waves, and stimulation-induced rhythmic or ictal discharges in β-lactam-induced neurotoxicity, particularly with cefepime (17). An alternative hypothesis is that our decoding methods may detect increased β-γ neural dynamics as a proxy for sharper EEG transients due to hyperexcitability.

Besides interacting with GABA receptors, β-lactam antibiotics might also affect N-methyl-D-aspartate (NMDA) receptors. This interaction could lead to EEG changes like those reported in anti-NMDA receptor encephalitis, where increased β neural dynamics have been observed (49,50). Some studies have suggested this effect in antibiotic-induced encephalopathies with psychotic features (19), but it has been primarily examined in the context of macrolides and quinolones. A possible mechanism involves β-lactam antibiotics activating the Glutamate transporter 1 (GLT1), thereby reducing synaptic glutamate levels and decreasing NMDA receptor activation. This process may lead to increased GLT1 expression and further reductions in glutamate concentrations (51–53). However, further research is required to better understand the connection between β-lactams, their neurological effects, and corresponding EEG patterns.

Finally, the absence of β-lactam exposure decoding for β neural dynamics in patients without neurological injury could be attributed to several factors. An underlying neurological pathology might disrupt the normal neural excitatory-inhibitory balance, increasing patients’ sensitivity to generating higher frequency oscillations and/or sharp transients observed in the beta band. Additionally, our limited dataset may have reduced statistical power in this subgroup, potentially contributing to the null result. Further studies are needed to investigate these possibilities.

We also observed a relationship between θ and α EEG activity and the SOFA score, which assesses the severity of organ failure in ICU patients. Θ and α EEG activity can reflect cerebral distress, a component of the SOFA score. Previous research has linked these neural dynamics to cerebral stress in ICU patients following open-heart surgery (54), mild traumatic brain injury (55), and stroke (56). Decreased variability or amplitude in α oscillations and changes in θ and α amplitude have been associated with poor prognosis in severe traumatic brain injury (57,58). Our results align with these findings, as the SOFA score’s neurological component specifically depends on the Glasgow coma scale score.

### Limitations

The method used in this study allowed us to find a relationship between β neural dynamics and β-lactam exposure. However, the number of patients and the diversity of β-lactam prescribed limited our ability to investigate the effect of any specific β-lactam on the neurophysiological data (Supplementary Figure 1). It would be valuable to refine our findings by focusing on β-lactams like cefepime, which are more likely to induce neurotoxicity (6,11,59). Moreover, these findings require validation in other patient cohorts before using them at the bedside. Unfortunately, we could not confirm clear β-lactam-induced neurotoxicity in the patients investigated here. Comparing the relationship between β-lactam exposure and EEG β neural dynamics with known neurophysiological patterns linked to β-lactam-induced neurotoxicity (6,17,19,60) could help define a neurotoxicity threshold, improving the monitoring of β-lactam treatment in critical care patients.

## Conclusions

In conclusion, our study suggested that the amplitude of neural dynamics within the β/low-γ frequency range (20-40 Hz) codes for β-lactam exposure in ICU patients, as revealed through multivariate decoding analysis on quantitative EEG. These findings, when combined with existing neurophysiological markers of β-lactam neurotoxicity, could improve the monitoring of these antibiotics in vulnerable populations such as those with acute brain injury or other organ dysfunction. However, further studies on larger cohorts are necessary to validate these results and incorporate them into clinical practice. This work underscores the potential of quantitative EEG and advanced multivariate decoding methods to identify subtle neurophysiological features that are otherwise difficult to detect.

## Data Availability

Numerical data supporting this study will be available on GitHub upon publication.

## List of abbreviations

EEG: Electroencephalography
qEEG: quantitative Electroencephalography
SOFA: Sepsis-related Organ Failure Assessment
ICU: Intensive Care Unit
TDM: Therapeutic Drug Monitoring
MIC: Minimum Inhibitory Concentration
GABA: Gamma Aminobutyric Acid
IPSP: Inhibitory Postsynaptic Potential
NMDA: N-methyl-D-aspartate
GLT1: Glutamate transporter 1

## Declarations

## Ethics approval and consent to participate

The current analysis was part of a prospective, multicenter trial conducted at two Intensive Care Units: Timone University Hospital and North University Hospital, Marseille (ClinicalTrials.gov ID NCT03339869. Registered 14 September 2017). The study complied with European data privacy regulations, as validated by the local data protection officer, and adhered to the principles outlined in the Declaration of Helsinki. The study received approval from the ethics committee *Comité de Protection des Personnes Sud-Est I* (Ref 2017-A01446-47).

## Consent for publication

Patients or their designated representatives were provided with clear, truthful, and relevant information, and consent was obtained prior to enrolment in this study.

## Availability of data and materials

Numerical data supporting this study will be available on GitHub upon publication.

## Authors’ contributions

Conceptualization: R.G., L.V, O.B. and A.Z. Methodology: A.Z., R.G and S.L. Investigation: L.V, M.L and A.Z. Visualization: A.Z. Funding acquisition: R.G, O.B, L.V and M.L. Project administration: R.G, S.L and A.Z. Supervision: R.G and S.L. Writing—original draft: A.Z. Writing—review and editing: all the authors.

## Acknowledgments

We thank Benjamin Morillon and Valentin Wyart for their valuable insights into this paper.

## Supplementary data

**Supplementary Figure 1.**
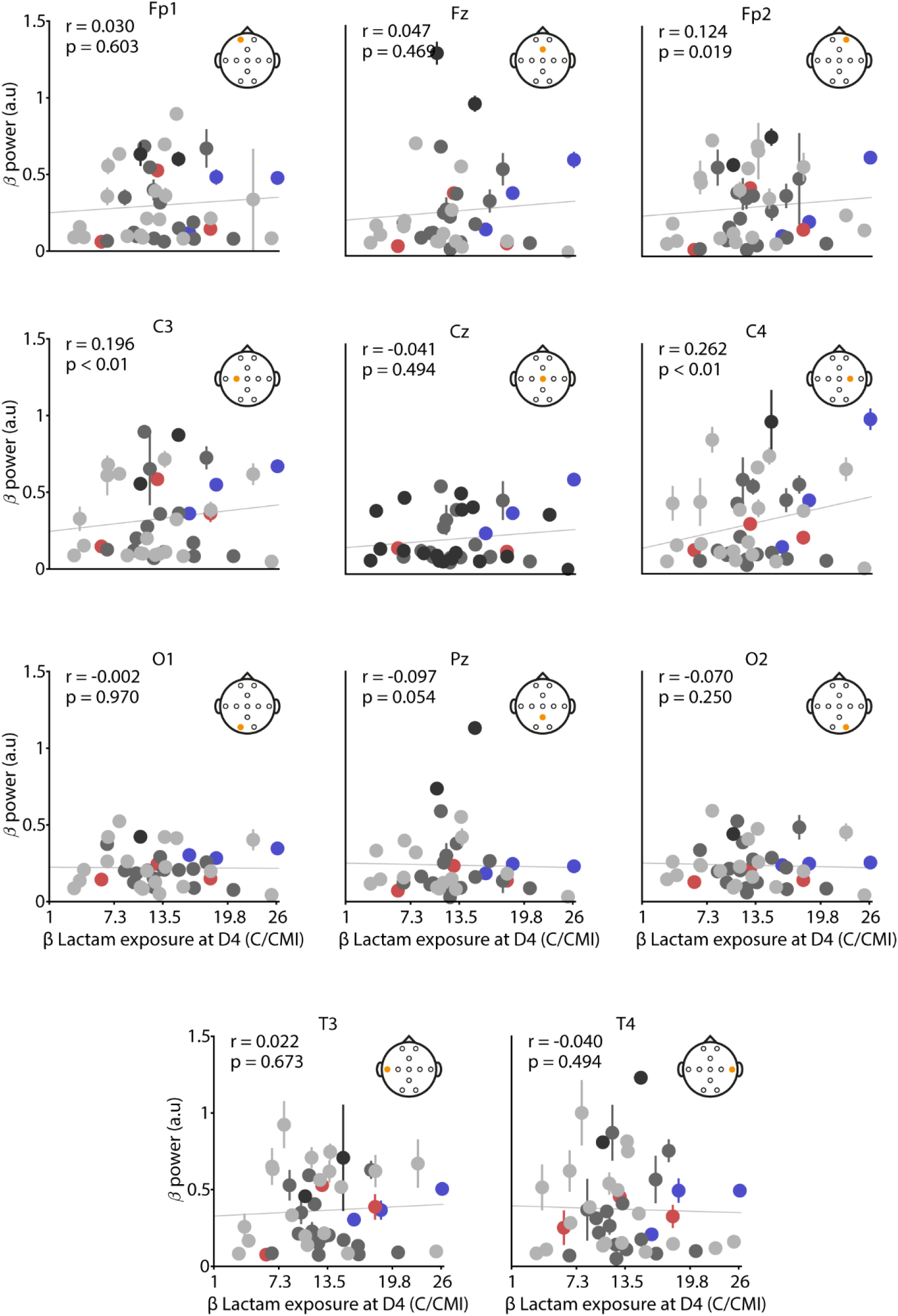
Correlation between the power of β neural dynamics for each EEG electrode and the β-lactam exposure. Each point relates to one patient. Light grey, dark grey, black, red and blue points correspond to the patients treated by piperacillin-tazobactam, cefotaxime, ceftazidime, meropenem and cefepime, respectively.

**Supplementary Figure 2.**
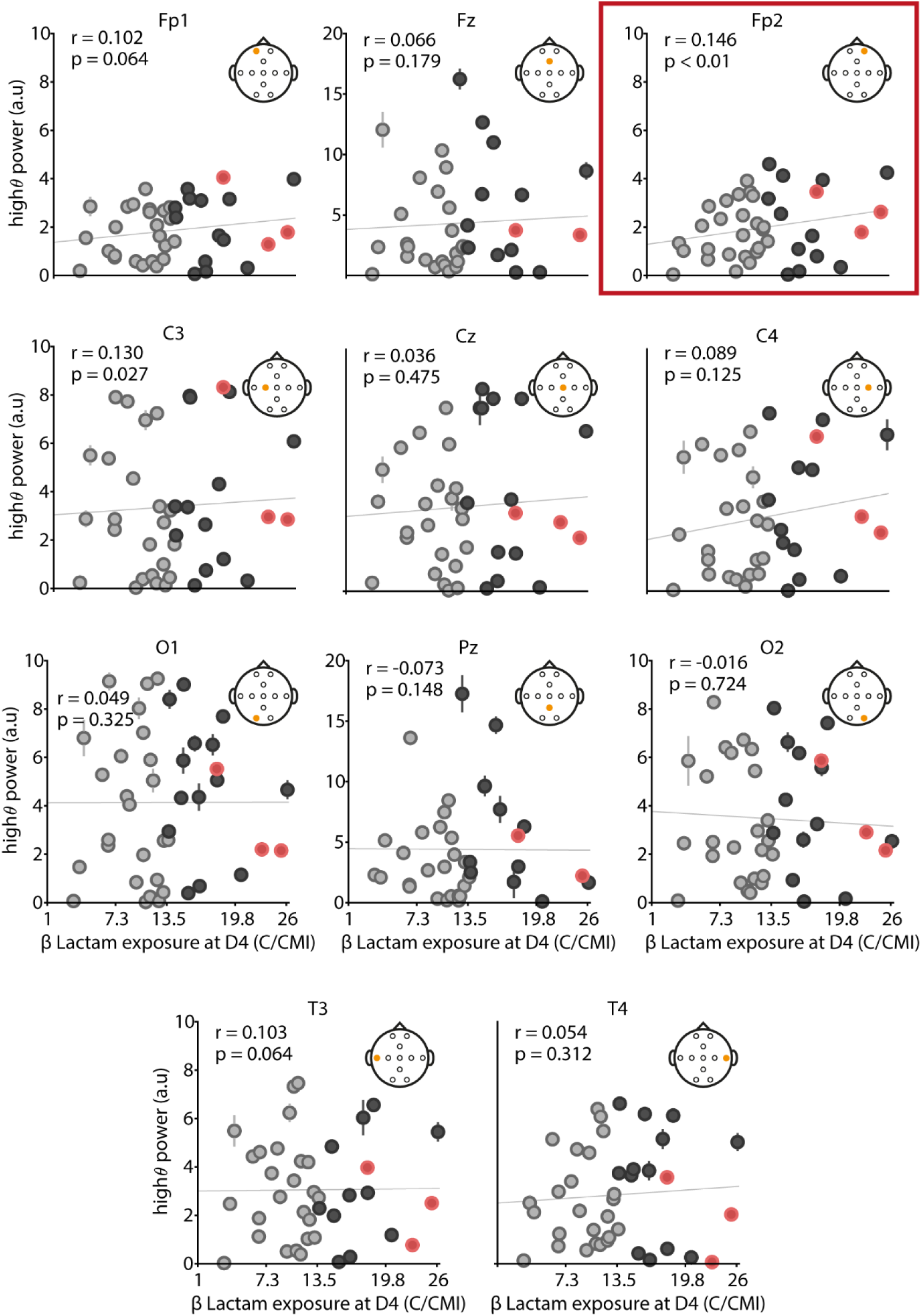
Correlation between the power of the high θ neural dynamics for each EEG electrode and the β-lactam exposure. Each point relates to one patient. Grey, black and red points correspond to the patients labelled as sub-, well and overdosed respectively. Electrodes outlined in red show a statistically significant positive correlation (p<0.01).

**Supplementary Figure 3.**
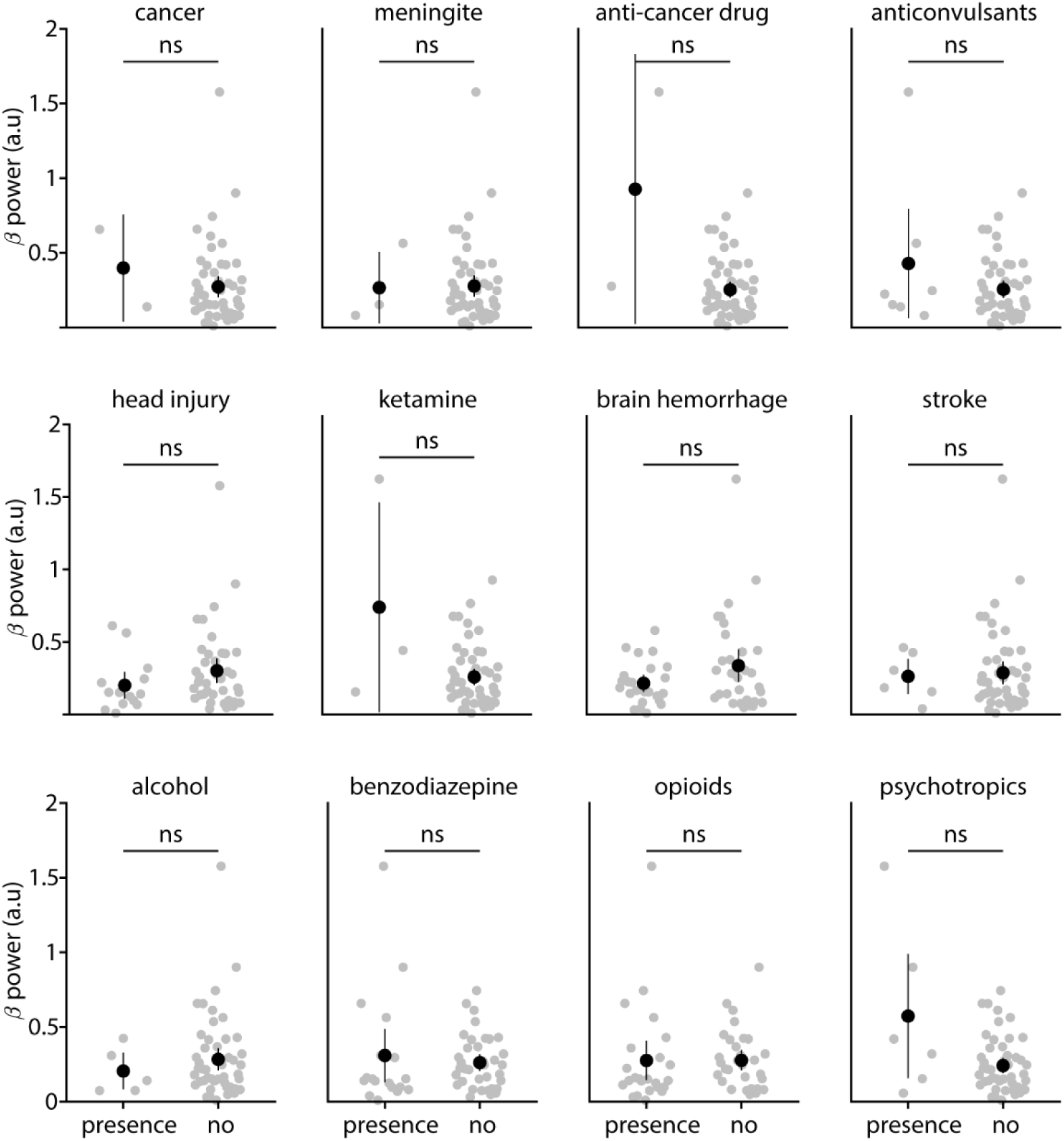
Comparison of the power of β neural dynamics between presence or absence of features found in the group. Not significant (ns) p > 0.05.

**Supplementary Figure 4.**
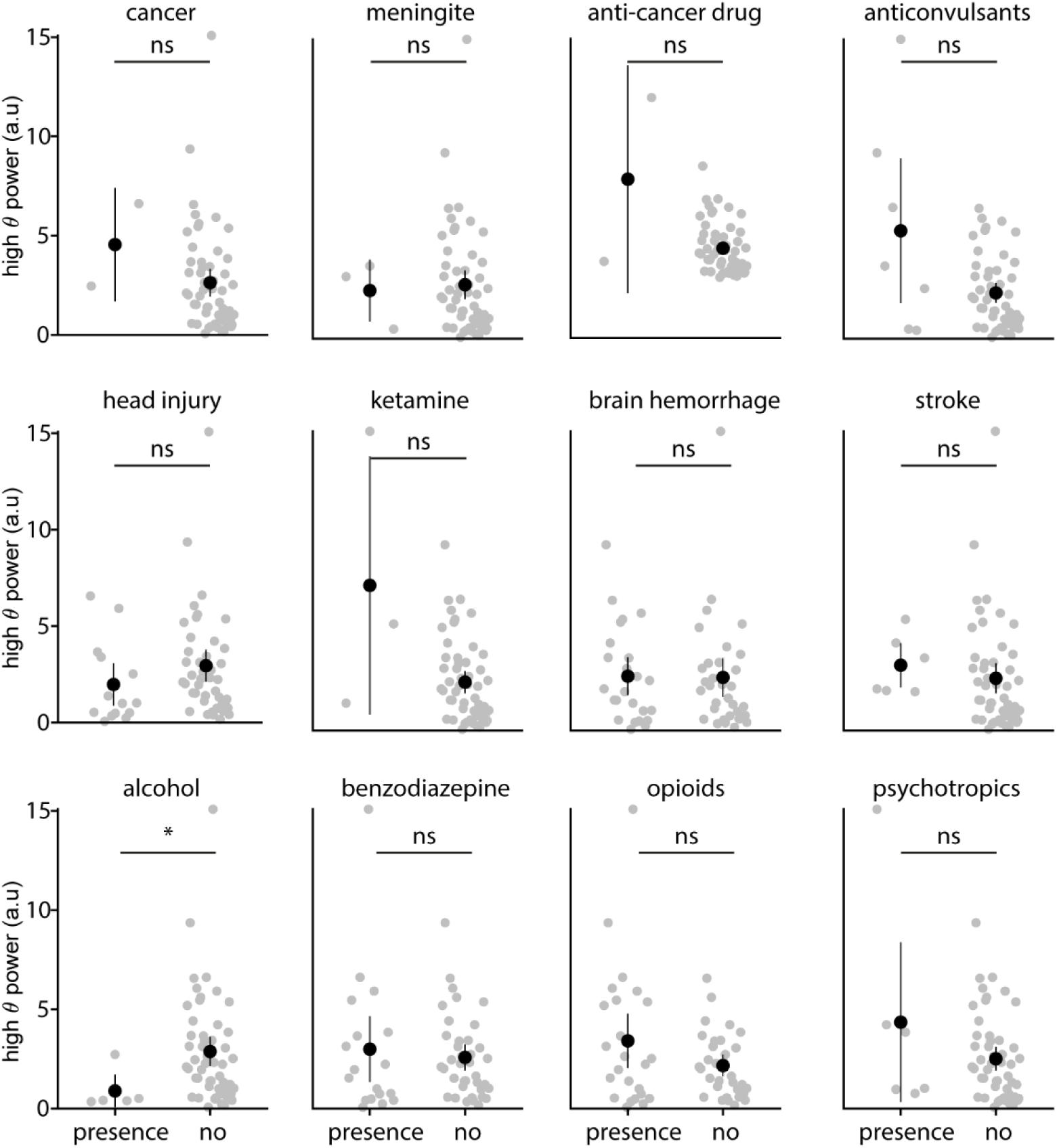
Comparison of the power of the high θ neural dynamics between presence or absence of features found in the group. Not significant (ns) p > 0.05; (*) p< 0.01.(Wilcoxon rank sum t-test).

**Supplementary Figure 5.**
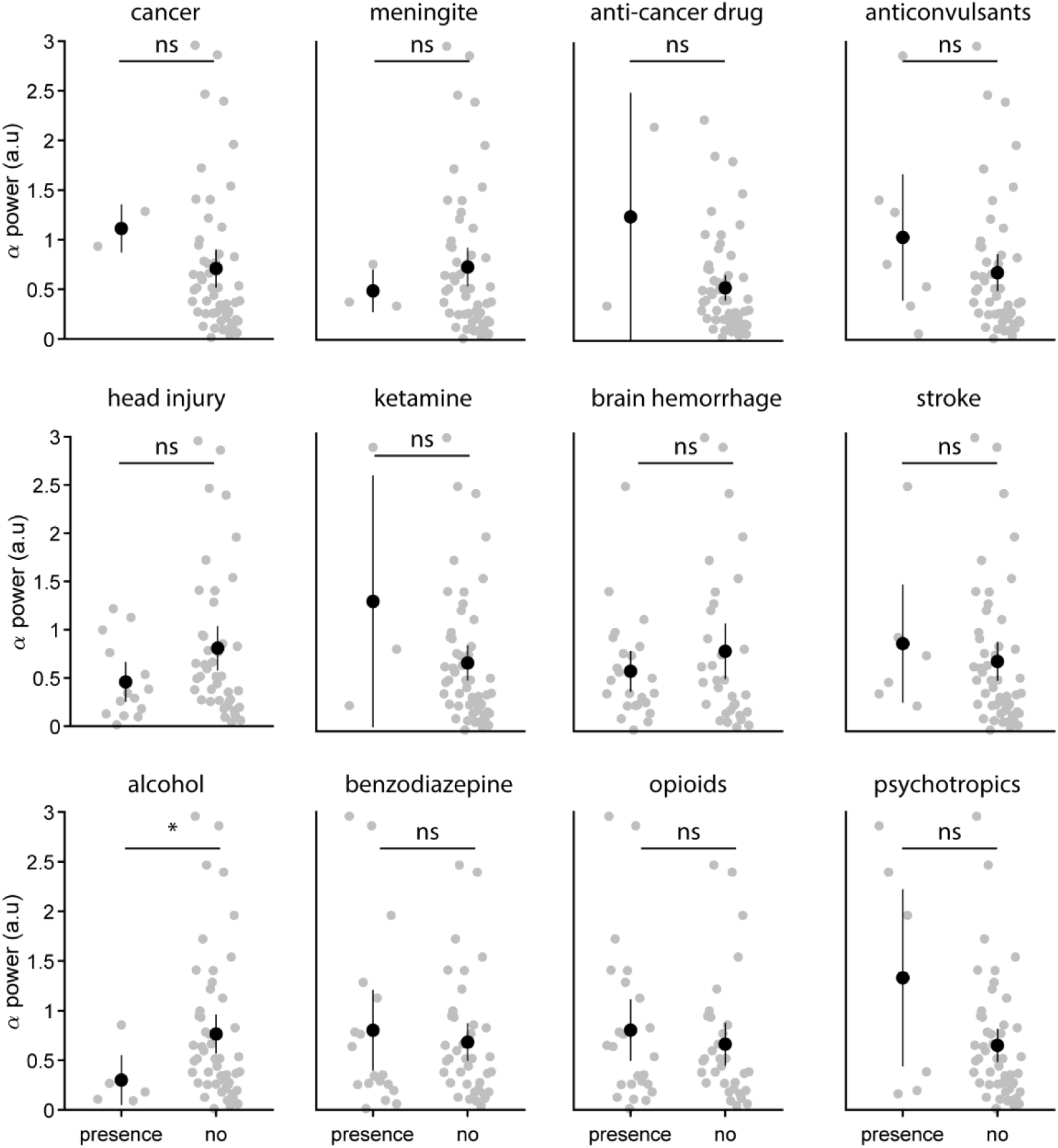
Comparison of the power of α neural dynamics between presence or absence of features found in the group. Not significant (ns) p > 0.05; (*) p< 0.01.(Wilcoxon rank sum t-test).

## References

1. Annual Epidemiological Report (europa.eu). Antimicrobial consumption in the EU / EEA (ESAC-Net). 2021;(November).

2. Baggs J, Fridkin SK, Pollack LA, Srinivasan A, Jernigan JA. Estimating national trends in inpatient antibiotic use among US hospitals from 2006 to 2012. JAMA Intern Med. 2016;176(11):1639–48.

3. Centers for Disease Control and Prevention. Antibiotic Use in the United States, 2017: Progress and Opportunities. Atlanta, GA US Dept Heal Hum Serv [Internet]. 2017;1–9. Available from: https://www.cdc.gov/antibiotic-use/stewardship-report/pdf/stewardship-report.pdf

4. Deshayes S, Coquerel A, Verdon R. Neurological Adverse Effects Attributable to β-Lactam Antibiotics: A Literature Review. Drug Saf. 2017;40(12):1171–98.

5. Blumenthal KG, Peter JG, Trubiano JA, Phillips EJ. Antibiotic allergy. Lancet [Internet]. 2019;393(10167):183–98. Available from: 10.1016/S0140-6736(18)32218-9

6. Lacroix C, Kheloufi F, Montastruc F, Bennis Y, Pizzoglio V, Micallef J. Serious central nervous system side effects of cephalosporins: A national analysis of serious reports registered in the French Pharmacovigilance Database. J Neurol Sci. 2019;398(December 2018):196–201.

7. Lacroix C, Bera-Jonville AP, Montastruc F, Velly L, Micallef J, Guilhaumou R. Serious neurological adverse events of ceftriaxone. Antibiotics. 2021;10(5):1–11.

8. Shenoy ES, Macy E, Rowe T, Blumenthal KG. Evaluation and Management of Penicillin Allergy: A Review. JAMA - J Am Med Assoc. 2019;321(2):188–99.

9. Vardakas KZ, Kalimeris GD, Triarides NA, Falagas ME. An update on adverse drug reactions related to β-lactam antibiotics. Expert Opin Drug Saf [Internet]. 2018;17(5):499–508. Available from: 10.1080/14740338.2018.1462334

10. Roger C, Louart B. Beta-lactams toxicity in the intensive care unit: An underestimated collateral damage? Microorganisms. 2021;9(7).

11. Payne LE, Gagnon DJ, Riker RR, Seder DB, Glisic EK, Morris JG, et al. Cefepime-induced neurotoxicity: A systematic review. Crit Care. 2017;21(1):1–8.

12. Fugate JE, Kalimullah EA, Hocker SE, Clark SL, Wijdicks EFM, Rabinstein AA. Cefepime neurotoxicity in the intensive care unit: A cause of severe, underappreciated encephalopathy. Crit Care. 2013;17(6).

13. Grahl JJ, Stollings JL, Rakhit S, Person AK, Wang L, Thompson JL, et al. Antimicrobial exposure and the risk of delirium in critically ill patients. Crit Care [Internet]. 2018 Dec 12 [cited 2024 Sep 7];22(1):1–8. Available from: https://ccforum.biomedcentral.com/articles/10.1186/s13054-018-2262-z

14. Guilhaumou R, Benaboud S, Bennis Y, Dahyot-Fizelier C, Dailly E, Gandia P, et al. Optimization of the treatment with beta-lactam antibiotics in critically ill patients. Crit Care [Internet]. 2019;23(1):104. Available from: https://ccforum.biomedcentral.com/articles/10.1186/s13054-019-2378-9

15. Gatti M, Cojutti PG, Bartoletti M, Tonetti T, Bianchini A, Ramirez S, et al. Expert clinical pharmacological advice may make an antimicrobial TDM program for emerging candidates more clinically useful in tailoring therapy of critically ill patients. Crit Care [Internet]. 2022 Dec 1 [cited 2024 Sep 7];26(1):1–14. Available from: https://ccforum.biomedcentral.com/articles/10.1186/s13054-022-04050-9

16. Roberts JA, Abdul-Aziz MH, Lipman J, Mouton JW, Vinks AA, Felton TW, et al. Individualised antibiotic dosing for patients who are critically ill: Challenges and potential solutions. Lancet Infect Dis. 2014;14(6):498–509.

17. Li HT, Lee CH, Wu T, Cheng MY, Tseng WEJ, Chang CW, et al. Clinical, Electroencephalographic Features and Prognostic Factors of Cefepime-Induced Neurotoxicity: A Retrospective Study. Neurocrit Care [Internet]. 2019;31(2):329–37. Available from: 10.1007/s12028-019-00682-y

18. Hirsch LJ, Fong MWK, Leitinger M, LaRoche SM, Beniczky S, Abend NS, et al. American Clinical Neurophysiology Society’s Standardized Critical Care EEG Terminology: 2021 Version. J Clin Neurophysiol. 2021;38(1):1–29.

19. Bhattacharyya S, Darby RR, Raibagkar P, Castro LNG, Berkowitz AL. Antibiotic-associated encephalopathy. Neurology. 2016;86(10):963–71.

20. Chow KM, Hui AC, Szeto CC. Neurotoxicity induced by beta-lactam antibiotics: From bench to bedside. Eur J Clin Microbiol Infect Dis. 2005;24(10):649–53.

21. Höller Y, Helmstaedter C, Lehnertz K. Quantitative Pharmaco-Electroencephalography in Antiepileptic Drug Research. CNS Drugs [Internet]. 2018 Sep 1 [cited 2024 Aug 21];32(9):839–48. Available from: https://link.springer.com/article/10.1007/s40263-018-0557-x

22. Canan S, Ankarali S, Marangoz C. Detailed spectral profile analysis of penicillin-induced epileptiform activity in anesthetized rats. Epilepsy Res. 2008;82:7–14.

23. Raposo J, Teotónio R, Bento C, Sales F. Amoxicillin, a potential epileptogenic drug. Epileptic Disord [Internet]. 2016 Dec 1 [cited 2024 Aug 15];18(4):454–7. Available from: https://pubmed.ncbi.nlm.nih.gov/27900944/

24. Silfverhuth MJ, Kortelainen J, Ruohonen J, Suominen K, Niinimäki J, Sonkajärvi E, et al. A characteristic time sequence of epileptic activity in EEG during dynamic penicillin-induced focal epilepsy--a preliminary study. Seizure [Internet]. 2011 Sep [cited 2024 Aug 15];20(7):513–9. Available from: https://pubmed.ncbi.nlm.nih.gov/21511498/

25. Legrand T, Vodovar D, Tournier N, Khoudour N, Hulin A. Simultaneous determination of eight β-lactam antibiotics, amoxicillin, cefazolin, cefepime, cefotaxime, ceftazidime, cloxacillin, oxacillin, and piperacillin, in human plasma by using ultra-high-performance liquid chromatography with ultraviolet detection. Antimicrob Agents Chemother. 2016;60(8):4734–42.

26. Verdier MC, Tribut O, Tattevin P, Tulzo Y Le, Michelet C, Bentué-Ferrer D. Simultaneous determination of 12 β-lactam antibiotics in human plasma by high-performance liquid chromatography with UV detection: Application to therapeutic drug monitoring. Antimicrob Agents Chemother. 2011;55(10):4873–9.

27. European Medicines Agency. ICH Guideline M10 on Bioanalytical Method Validation. 2019;44(March):57.

28. Vincent J-L, Moreno R, Takala J, Willatts S, De Mendon∼a A, Bruining H, et al. The SOFA (Sepsis.related Organ Failure Assessment) score to describe organ dysfunction/failure. Intensive Care Med. 1996;22:707–10.

29. Seeck M, Koessler L, Bast T, Leijten F, Michel C, Baumgartner C, et al. The standardized EEG electrode array of the IFCN. Clin Neurophysiol. 2017 Oct 1;128(10):2070–7.

30. Oostenveld R, Fries P, Maris E, Schoffelen J-M. FieldTrip: Open Source Software for Advanced Analysis of MEG, EEG, and Invasive Electrophysiological Data. Comput Intell Neurosci [Internet]. 2011 [cited 2024 May 3];2011. Available from: http://www.mathworks.com

31. Varoquaux G, Raamana PR, Engemann DA, Hoyos-Idrobo A, Schwartz Y, Thirion B. Assessing and tuning brain decoders: Cross-validation, caveats, and guidelines. Neuroimage [Internet]. 2017;145(August 2015):166–79. Available from: 10.1016/j.neuroimage.2016.10.038

32. Storey JD. A direct approach to false discovery rates. J R Stat Soc Ser B (Statistical Methodol [Internet]. 2002 Aug 1 [cited 2024 Apr 24];64(3):479–98. Available from: https://onlinelibrary.wiley.com/doi/full/10.1111/1467-9868.00346

33. Moriyama H, Nomura S, Kida H, Inoue T, Imoto H, Maruta Y, et al. Suppressive effects of cooling compounds icilin on penicillin G-induced epileptiform discharges in anesthetized rats. Front Pharmacol [Internet]. 2019 Jun 13 [cited 2024 Jul 13];10(JUN):437537. Available from: www.frontiersin.org

34. Porjesz B, Almasy L, Edenberg HJ, Wang K, Chorlian DB, Foroud T, et al. Linkage disequilibrium between the beta frequency of the human EEG and a GABAA receptor gene locus. Proc Natl Acad Sci U S A. 2002;99(6):3729–33.

35. Whittington MA, Traub RD, Faulkner HJ, Stanford IM, Jeffers JGR. Recurrent excitatory postsynaptic potentials induced by synchronized fast cortical oscillations. Proc Natl Acad Sci U S A. 1997;94(22):12198–203.

36. DeLorey TM, Olsen RW. γ-Aminobutyric acid(A) receptor structure and function. J Biol Chem. 1992;267(24):16747–50.

37. Wallace KL. Antibiotic-induced convulsions. Crit Care Clin. 1997;13(4):741–62.

38. Grill MF, Maganti R. Cephalosporin-induced neurotoxicity: Clinical manifestations, potential pathogenic mechanisms, and the role of electroencephalographs monitoring. Ann Pharmacother. 2008;42(12):1843–50.

39. Lindquist CEL, Dalziel JE, Cromer BA, Birnir B. Penicillin blocks human α1β1 and α1β1γ2S GABAA channels that open spontaneously. Eur J Pharmacol [Internet]. 2004 [cited 2024 Aug 12];496(1–3):23–32. Available from: www.elsevier.com/locate/ejphar

40. Yamazaki S, Mochizuki Y, Terai T, Sugimoto M, Uchida I, Matsuoka N, et al. Intracerebroventricular injection of the antibiotic cefoselis produces convulsion in mice via inhibition of GABA receptors. Pharmacol Biochem Behav. 2002;74(1):53–9.

41. Davidoff RA. Penicillin and inhibition in the cat spinal cord. Brain Res. 1972 Oct 27;45(2):638–42.

42. Dingledine R, Gjerstad L. Penicillin blocks hippocampal IPSPs, unmasking prolonged EPSPs. Brain Res. 1979 May 18;168(1):205–9.

43. van Duijn H, Schwartzkroin PA, Prince DA. Action of penicillin on inhibitory processes in the cat’s cortex. Brain Res. 1973 Apr 27;53(2):470–6.

44. Meyer H, Prince D. Convulsant actions of penicillin: effects on inhibitory mechanisms. Brain Res. 1973 Apr 27;53(2):477–82.

45. Wong RKS, Prince DA. Dendritic Mechanisms Underlying Penicillin-Induced Epileptiform Activity. Science (80-) [Internet]. 1979 [cited 2024 Aug 12];204(4398):1228–31. Available from: https://www.science.org/doi/10.1126/science.451569

46. Villalobos N, Magdaleno-Madrigal VM. Pallidal GABA B receptors: involvement in cortex beta dynamics and thalamic reticular nucleus activity. J Physiol Sci [Internet]. 2023 Jun 16 [cited 2024 Aug 12];73(1):14. Available from: https://jps.biomedcentral.com/articles/10.1186/s12576-023-00870-8

47. Visser SAG, Wolters FLC, Gubbens-Stibbe JM, Tukker E, Van Der Graaf PH, Peletier LA, et al. Mechanism-Based Pharmacokinetic/Pharmacodynamic Modeling of the Electroencephalogram Effects of GABAA Receptor Modulators: In Vitro-in Vivo Correlations. J Pharmacol Exp Ther [Internet]. 2003 Jan 1 [cited 2024 Aug 12];304(1):88–101. Available from: https://jpet.aspetjournals.org/content/304/1/88

48. Viloria-Alebesque A, Povar-Echeverría M, Bruscas-Alijarde MJ, Gracia-Gutiérrez A, Royo-Trallero L, Al-Cheikh-Felices P. Myoclonus induced by amoxicillin-clavulanic acid. Epilepsy Behav Reports [Internet]. 2020;14:100367. Available from: 10.1016/j.ebr.2020.100367

49. Jeannin-Mayer S, André-Obadia N, Rosenberg S, Boutet C, Honnorat J, Antoine JC, et al. EEG analysis in anti-NMDA receptor encephalitis: Description of typical patterns Anti-NMDAR Encephalitis EEG patterns Extreme delta brush Generalized rhythmic delta activity Excessive beta activity h i g h l i g h t s. Clin Neurophysiol [Internet]. 2019 [cited 2024 Jul 13];(130):289–96. Available from: 10.1016/j.clinph.2018.10.017

50. Zhang Y, Liu G, Di Jiang M, Ping Li L, Ying Su Y. Analysis of electroencephalogram characteristics of anti-NMDA receptor encephalitis patients in China. Clin Neurophysiol [Internet]. 2017 [cited 2024 Jul 13];(128):1227–33. Available from: 10.1016/j.clinph.2017.04.015

51. Abulseoud OA, Alasmari F, Hussein AM, Sari Y. Ceftriaxone as a Novel Therapeutic Agent for Hyperglutamatergic States: Bridging the Gap Between Preclinical Results and Clinical Translation. Front Neurosci [Internet]. 2022 Jul 5 [cited 2024 Jul 13];16:841036. Available from: www.frontiersin.org

52. Rothstein JD, Patel S, Regan MR, Haenggeli C, Huang YH, Bergles DE, et al. β-Lactam antibiotics offer neuroprotection by increasing glutamate transporter expression. Nature [Internet]. 2005 Jan 6 [cited 2024 Jul 13];433(7021):73–7. Available from: https://www.nature.com/articles/nature03180

53. Smaga I, Fierro D, Mesa J, Filip M, Knackstedt LA. Molecular changes evoked by the beta-lactam antibiotic ceftriaxone across rodent models of substance use disorder and neurological disease. Neurosci Biobehav Rev [Internet]. 2020 [cited 2024 Jul 13];(115):116–30. Available from: 10.1016/j.neubiorev.2020.05.016

54. Plaschke K, Fichtenkamm P, Schramm C, Hauth S, Martin E, Verch M, et al. Early postoperative delirium after open-heart cardiac surgery is associated with decreased bispectral EEG and increased cortisol and interleukin-6. Intensive Care Med. 2010;36(12):2081–9.

55. Kaltiainen H, Helle L, Liljeström M, Renvall H, Forss N. Theta-Band Oscillations as an Indicator of Mild Traumatic Brain Injury. Brain Topogr [Internet]. 2018;31(6):1037–46. Available from: 10.1007/s10548-018-0667-2

56. Vivaldi N, Caiola M, Solarana K, Ye M. Evaluating Performance of EEG Data-Driven Machine Learning for Traumatic Brain Injury Classification. IEEE Trans Biomed Eng. 2021;68(11):3205–16.

57. Tolonen A, Särkelä MOK, Takala RSK, Katila A, Frantzén J, Posti JP, et al. Quantitative EEG parameters for prediction of outcome in severe traumatic brain injury: Development study. Clin EEG Neurosci. 2018;49(4):248–57.

58. Vespa PM, Boscardin WJ, Hovda DA, Mcarthur DL, Nuwer MR, Martin NA, et al. Early and persistent impaired percent alpha variability on continuous electroencephalography monitoring as predictive of poor outcome after traumatic brain injury. J Neurosurg. 2002;97:84–92.

59. Triplett JD, Lawn ND, Chan J, Dunne JW. Cephalosporin-related neurotoxicity: Metabolic encephalopathy or non-convulsive status epilepticus? J Clin Neurosci [Internet]. 2019;67(April 2018):163–6. Available from: 10.1016/j.jocn.2019.05.035

60. Grill MF, Maganti RK. Neurotoxic effects associated with antibiotic use: Management considerations. Br J Clin Pharmacol. 2011;72(3):381–93.

